# Promising efficacy of following a third dose of mRNA SARS-CoV-2 vaccination in patients treated with anti-CD20 antibody who failed 2-dose vaccination

**DOI:** 10.1101/2022.04.28.22274174

**Authors:** Yohei Funakoshi, Kimikazu Yakushijin, Goh Ohji, Wataru Hojo, Hironori Sakai, Marika Watanabe, Akihito Kitao, Yoshiharu Miyata, Yasuyuki Saito, Shinichiro Kawamoto, Katsuya Yamamoto, Mitsuhiro Ito, Taiji Koyama, Yoshinori Imamura, Naomi Kiyota, Hiroshi Matsuoka, Yasuko Mori, Hironobu Minami

## Abstract

Anti-CD20 antibodies react with CD20 expressed not only on malignant B cells but also on normal B cells. It has been reported that patients treated with anti-CD20 antibodies had an insufficient response to two-dose mRNA SARS-CoV-2 vaccination. To investigate the efficacy of a third dose in these patients, we investigated serum IgG antibody titers for S1 protein after third vaccination in 22 patients treated with anti-CD20 antibody who failed two-dose vaccination. Results showed that overall, 50% of patients seroconverted. Although no patient who received the third dose within 1 year of the last anti-CD20 antibody administration showed an increase in S1 antibody titer, 69% of patients who received the third dose more than 1 year after the last anti-CD20 antibody administration seroconverted. Our data show that a third dose of vaccination is effective in improving seroconversion rate in patients treated with anti-CD20 antibody who failed standard two-dose vaccination.

## Research letter

Patients with hematological malignancies, especially those receiving anti-CD20 antibody therapy, have increased morbidity and mortality from the coronavirus disease 2019 (COVID-19) infection [1]. Because the anti-CD20 antibodies rituximab and obinutuzumab react with CD20 expressed not only on malignant B cells but also on normal B cells, they impair the efficacy of SARS-CoV-2 mRNA vaccination in triggering the humoral immune response. In reality, it has been reported that patients treated with anti-CD20 antibodies had an insufficient response after two-dose vaccination compared with an age-matched healthy cohort, and a third dose (booster) vaccination in these patients is accordingly expected to improve immunogenicity [2]. However, data for third-dose vaccination remain insufficient. In our previous study, we investigated S1 antibody titers 2 weeks after the second dose of mRNA SARS-CoV-2 vaccination, BNT162b2, in patients with B cell malignancy treated with anti-CD20 antibody before vaccination [3]. Results showed that many patients (29 of 39) failed to achieve seroconversion (the optimal optical density (O.D.) cut-off value of anti-S1 IgG antibody for seroconversion was determined to be 0.26.) [3, 4]. In the present study, we investigated the efficacy of a third dose of vaccination in those patients in our previous study in whom the second dose failed to produce seroconversion.

Twenty two of 29 patients who failed to seroconvert after second-dose vaccination in our previous study provided written informed consent to participate in this study. The protocol was approved by Kobe University Hospital Ethics Committee (No. B2056714, 1481). All patients (median age 74 years, range 57–86) received a third dose of vaccination (BNT162b2, n = 18 and mRNA-1273, n = 4, respectively) 6–50 (median 18.5) months after the final dose of anti-CD20 antibody (rituximab, n = 21 and obinutuzumab, n = 1). Diagnoses included diffuse large B-cell lymphoma (n = 13), follicular lymphoma (n = 6), lymphoplasmacytic lymphoma/Waldenström’s macroglobulinemia (n = 2), and mantle cell lymphoma (n = 1). No patient had received any chemotherapy after the last anti-CD20 antibody dose. S1 antibody titers were measured approximately 14 days after the third-dose vaccination (median 13 days, range 7–17) with a COVID-19 Human IgM IgG ELISA kit (Spike Protein-S1) (Cellspect, Inc., Japan). Results showed that overall, 50% (11/22) of patients seroconverted (Figure 1A). Importantly, no patient who received the third dose of vaccination within 1 year of the last anti-CD20 antibody administration showed an increase in S1 antibody titer (0/6, Figure 1B). On the contrary, 69% patients (11/16) who received the third dose of vaccination more than 1 year after the last anti-CD20 antibody administration seroconverted. Finally, we investigated surrogate markers of antibody production ability. We found no relationship between the percentage of peripheral blood B-cells (CD19-positive cells) or serum IgG level and S1 antibody titer (r^2^ = 0.10 and 0.14, respectively; Figure 1C and D).

**Figure 1.**
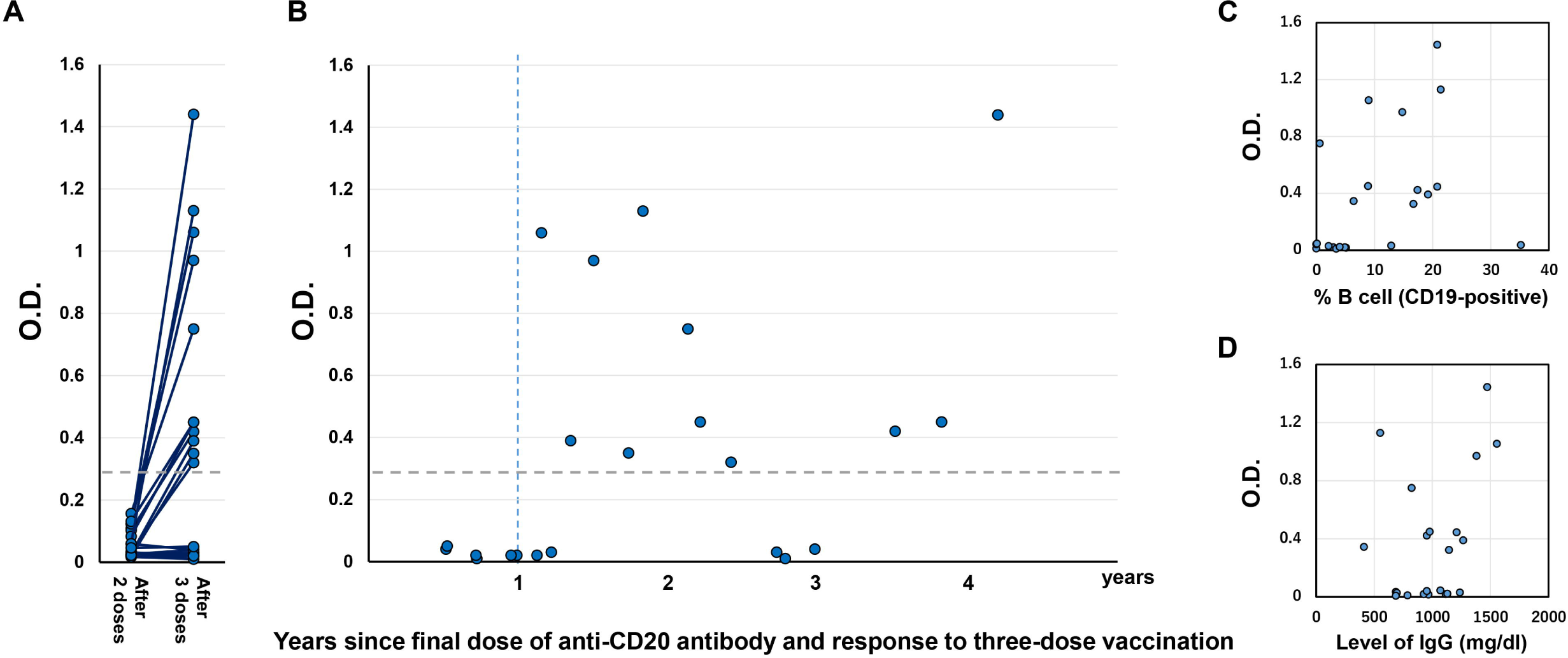
(A) Humoral quantitative anti-spike 1 (S1) antibody response 14 days (± 7 days) after the second and third vaccination in B-cell lymphoma patients treated with anti-CD20 antibody (n = 22). The horizontal dotted line indicates the threshold (0.26) for seroconversion. (B) Relationship between S1 titer and time since from the final dose of anti-CD20 antibody to the third dose of vaccination. The horizontal dotted line indicates the threshold (0.26) for seroconversion. (D) Correlation between S1 antibody titer and percentage of CD19-positive B cells (normal range 6–23%). (E) Correlation between S1 antibody titer and total IgG level (normal range 870–1700 mg/dL).

In this study, to test the efficacy of a third dose of vaccination in patients treated with anti-CD20 antibody, we enrolled only non-seroconverted patients after the second dose of vaccination. Although there was concern that the intense immunosuppression caused by anti-CD20 antibodies could persist over the long term, most patients (69%) who received the third dose of vaccine more than 1 year after the last anti-CD20 antibody administration seroconverted. This result shows that mRNA vaccination is indicated even in patients who have received B cell depletion therapy. Nevertheless, there are some issues concerning the booster shot. First, seroconversion following the third dose was not seen in patients within one year from the final dose of anti-CD20 antibody. Second, about 30% of patients did not achieve seroconversion even more than 1 year after the final dose of anti-CD20 antibody. Furthermore, seroconversion rate did not appear to increase with increasing years after the final of dose of anti-CD20 therapy. We previously reported that an increase in antibody titers following two-dose vaccination remained limited for a long time (more than 3 years) after the final dose of anti-CD20 antibody [3]. These results are consistent with the idea that long-term immunosuppression is maintained. In these patients, an alternative protection method might be necessary.

In conclusion, our data show that the third dose of vaccination is effective in improving seroconversion rate in patients treated with anti-CD20 antibody who failed standard two-dose vaccination.

## Data Availability

All data produced in the present study are available upon reasonable request to the authors.

